# Whole-exome sequencing in 415,422 individuals identifies rare variants associated with mitochondrial DNA copy number

**DOI:** 10.1101/2022.06.22.22276774

**Authors:** Vamsee Pillalamarri, Wen Shi, Conrad Say, Stephanie Yang, John Lane, Eliseo Guallar, Nathan Pankratz, Dan E. Arking

## Abstract

Inter-individual variation in the number of copies of the mitochondrial genome, termed mitochondrial DNA copy number (mtDNA-CN), reflects mitochondrial function and has been associated with various aging-related diseases. We examined 415,422 exomes of self-reported White ancestry individuals from the UK Biobank and tested the impact of rare variants, both at the level of single variants and through aggregate variant-set tests, on mtDNA-CN. A survey across nine variant sets tested enrichment of putatively causal variants and identified 14 genes at experiment-wide significance and three genes at marginal significance. These included associations at known mitochondrial DNA depletion syndrome genes (mtDNA helicase *TWNK*, p=5.7×10^−29^; mitochondrial transcription factor *TFAM*, p=4.3×10^−13^; mtDNA maintenance exonuclease *MGME1*, p=1.3×10^−6^) and the V617F dominant gain-of-function mutation in the tyrosine kinase *JAK2* (p=7.1×10^−17^) associated with myeloproliferative disease. Novel genes included the ATP-dependent protease *CLPX* (p=9.9×10^−9^) involved with mitochondrial proteome quality and the mitochondrial adenylate kinase *AK2* (p=5.3×10^−8^) involved with hematopoiesis. The most significant association was a missense variant in *SAMHD1* (p=4.2×10^−28^), found on a rare, 1.2 Mb shared ancestral haplotype on chromosome 20. *SAMHD1* encodes a cytoplasmic host restriction factor involved with viral defense response and the mitochondrial nucleotide salvage pathway, and is associated with Aicardi-Goutières syndrome 5, a childhood encephalopathy and chronic inflammatory response disorder. Rare variants were enriched in Mendelian mtDNA depletion syndrome loci, and these variants further implicated core processes in mtDNA replication, nucleoid structure formation, and maintenance. Together, these data indicate strong-effect mutations from the nuclear genome contribute to the genetic architecture of mtDNA-CN.

## Introduction

Mitochondria play vital roles in meeting the chemical energy requirements for the cell through oxidative phosphorylation (OXPHOS), in intrinsic apoptosis activation^1^, and in the regulation of innate immunity^2,3^. Most of mitochondrial form and function, including factors required to construct the organelle^4^, is encoded within the nuclear genome, yet mitochondria have evolved^5,6^ to retain a haploid circular, double-stranded, intron-free ∼ 16.6 kb DNA molecule (mtDNA) that encodes 13 proteins required for cellular respiration, including most of the components of OXPHOS, and 24 RNAs required for mitochondrial translational machinery. Thus, defects in human mitochondrial genomes, both at levels considered heteroplasmic and homoplasmic, contribute to disease risk^7^ including Mendelian mtDNA depletion syndromes^8^ and aging-related disease^9^.

Unlike the fixed content of nuclear DNA, mtDNA ploidy is variable, both within cells and across tissues, individuals, and cohorts^10,11^. Indeed, a portion of inter-individual variation in the number of mitochondrial genome copies (mtDNA-CN) reflects mitochondrial function and is heritable under nuclear genetic control via common polygenic variation^12,13^. As a minimally invasive biomarker, mtDNA-CN is associated with several aging-relevant traits including cardiovascular disease^11,14–16^, frailty and all-cause mortality^17^, neurodegeneration^18–20^, and cancer^21,22^.

Genome-wide association studies (GWAS) for mtDNA-CN have revealed mtDNA-CN to be a complex phenotype, with SNP-based heritability estimates between 7-8%. Family-based studies, however, have estimated broad sense heritability for mtDNA-CN at 54-65%^23,24^, and variance explained by common variants in a non-discovery cohort was 1-2%^12^. Rare genetic variants, especially mutations within protein-coding regions of the nuclear genome, collectively account for a substantial proportion of missing narrow-sense heritability for complex traits^25^. We therefore hypothesized that rare nuclear variants drive a significant proportion of variation in mtDNA-CN. Herein, we conduct a biobank-scale exome-based study of blood buffy-coat-derived mtDNA-CN in 415,422 individuals of European ancestry.

## Materials and Methods

### Sample and variant selection

Data for this study were obtained from the UK Biobank^26^, a resource containing dense genomics, lifestyle, and phenotyping data on ∼500,000 volunteers between the ages of 40 – 69 at recruitment, selectively invited to participate in the prospective study based on the NHS register and residing within a traveling distance to one of 22 assessment centers in the United Kingdom (UK). Blood collection for blood cell counts and genomics source material extraction was conducted during the recruitment period from 2006 – 2010. Genotype calls in PLINK format of whole exome sequencing (WES) data for 454,756 samples were accessed on the UK Biobank Research Access Platform (RAP) under application 17731. We started with a total of 26,645,252 variants across autosomal and chromosome X (including targeted and off-target) regions on GRCh38. The samples were further processed to remove individuals marked as having withdrawn research consent (45 samples removed), those of self-reported non-White race (25,754 removed using data field 21000, retaining any self-report of White British, White Irish, or any other white background) and those without a computable mtDNA-CN phenotype (13,535 removed) for a total of 415,422 samples.

### Computation of mtDNA-CN

mtDNA-CN was estimated from DNA derived from buffy coat, using a combination of whole exome and genotyping array data as previously described^12^. In brief, we first extracted reads from 49,997 UKB exome sequencing libraries (July 2018 release) and modeled the number of mapped sequencing reads to the reference GRCh38 ‘chrMT’ sequence as a function of total mapped reads, reads to unknown contigs, and decoy sequence reads. Nonlinear effects of total, unknown, and decoy read counts were incorporated using natural splines. The residuals of the adjusted mtDNA mapped read count metric were then regressed onto mitochondrial genotyping probe intensities, available for all 500k UKB participants^26^, in separate models for the UK BiLEVE (n=181 probes) and Affymetrix Axiom (n=244 probes) genotyping arrays, including in each model nuclear probe intensity-derived principal components to account for technical artifacts. The betas from these models were used to generate fitted values of the mtDNA read count metric in the entire exome sequencing cohort used in this study. Finally, we adjusted the metric for age, sex, and blood cell counts^27^ (leukocytes, red blood cells, nucleated red blood cells, platelets, lymphocytes, neutrophils, eosinophils, and basophils). All analyses utilized a standardized metric with mean 0 and standard deviation 1. mtDNA-CN was not computed for 13,535 samples that had missing probe intensities or had cell count outliers.

### Power Analysis

We computed the power of the study to detect rare variant associations along the spectrum of expected minor allele frequency (MAF) and effect sizes for hypothesized associations to mtDNA-CN. We based this prediction of the genetic architecture of mtDNA-CN on 129 previously identified genome-wide independent common variants associated with mtDNA-CN^12^, extrapolating a nonlinear power-law regression model fit of MAF and effect sizes of the independent common markers (Supplemental Figure S1). The R package genpwr version 1.0.4^28^ was used for power calculations.

### Single variant analysis

Single-variant association testing was implemented using the BOLT-LMM mixed-model association testing algorithm^29^ forcing the use of a non-infinitesimal mixed-model association test. BOLT-LMM computes a genetic relationship matrix (GRM, or ‘kinship matrix’) internally given a genetic marker set that spans the whole genome. Thus, to account for random effects of genetic relatedness within the BOLT-LMM mixed-model framework, we merged the exome-based genotype calls with 784,256 array-based genotyping calls from the same 415,422 individuals after lifting the coordinates for the array-based variants to hg38 from hg19, then provided a list of 672,036 (85.7%) of the array-based SNPs, restricted to MAF ≥ 0.001 and SNP-missingness ≤ 10%, to allow for construction of the GRM. The merged variants were filtered for variant- and sample-level missingness > 10% (1,193,215 variants removed), monomorphic variants (i.e., those with MAF = 0; 3,286,025 variants removed), and variants with minor allele count < 6 (19,720,859 variants removed), for a total analysis set of 6,766,699 variants (26.0% of all genotyped exome variants) across autosomes and chrX.

Covariates for analyses included age, sex, and 40 principal components (PCs) capturing genetic ancestry, and WES batch (45,200 from 50k initial exomes release and 370,222 from the remaining exomes released). WES batch effects were present due to the use of different oligo lots for exome probe capture in the first tranche. In addition, genotypes at 129 independent common autosomal variants and four independent common chrX variants, each previously associated with mtDNA-CN^12^, were included as covariates for autosomal and chrX variant testing, respectively. mtDNA-CN was then regressed onto the genotype using a linear mixed model, including the covariates as fixed effects and a GRM for random effects. For chrX, association testing was stratified by sex, and test statistics and standard errors from the analysis of each sex were combined with METAL^30^. BOLT-LMM internally removed an additional 757 individuals without all covariates available, yielding an analysis set of 414,665 individuals.

### Aggregate variant analysis

Aggregate rare variant testing was performed within a mixed-model association testing framework as implemented in the R Bioconductor package GENESIS^31^ v2.21.4. We restricted the analysis to 24,949,430 (95.9%) variants at ≤ 1% MAF across all exome-captured regions. A null mixed model was first fit for mtDNA-CN by regressing the phenotype onto fixed effect covariates of age, sex, genotyping PCs 1-40, and WES batch, and including a sparse GRM to account for random effects due to genetic relatedness. The sparse GRM was constructed from kinship estimates provided by the UK Biobank, setting all pairwise relationships with kinship coefficients < 0.0442 (3rd degree or less) to 0. The previously discovered common SNPs were not included as covariates for aggregate testing due to computational cost. Next, for each protein coding gene as defined in GENCODE v38^32^ corresponding to GRCh38.p13, rare-variant burden^33^ and adjusted, asymptotically independent sequence kernel association test (SKAT^34^) p-values were computed then combined within a single set-based mixed model association test (SMMAT^35^) to produce a single p-value from a chi-squared distribution with 4 degrees of freedom (df). Multiple SMMAT tests were carried out for each gene with overlapping sets of rare variants chosen to test various levels of enrichment of putatively causal variants. Specifically, for each protein-coding gene, a SMMAT test was run for nonsynonymous coding sequence variants passing increasing mutational severity thresholds (all nonsynonymous variants, nonsynonymous variants with CADD-PHRED^36^ score ≥ 18, predicted loss-of-function (pLoF) variants), and decreasing MAF (≤1%, ≤0.1%, ≤0.01%), such that in total we tested 9 combinations of rare variants assessing various levels of deleteriousness and allele frequency (Supplemental Table S1). Each test was required to have a minimum gene-level cumulative allele frequency of 0.01%, but no minimum minor allele count cutoff was set such that singletons and ultra-rare variants with MAC < 6 were included within the aggregate set. This yielded 153,481 total SMMAT tests. We reasoned that since the rare variants chosen across different sets for a given gene will have numerous variants in common, the test statistic p-values will also be correlated. A correlation matrix of the p-values for 13,581 common genes tested across all nine variant sets was used to perform hierarchical clustering that we conservatively interpreted as outlining four clusters of related p-values (Supplemental Figure S2). Therefore, the number of effective tests was set to the number of unique tested protein coding genes times four, and a correlation-based test p-value cutoff was set to 0.05 / (18557 × 4).

### Gene-level iterative leave-one-out and conditional variant analyses

For each study-wide significant gene set signal, we used both leave-one-out and gene level conditional analysis to assess the independence of rare variants at that locus. Leave-one-out analysis is a qualitative method that consists of iteratively dropping one variant from a given gene’s variant set and re-computing a SMMAT p-value for the locus, producing a measure of the relative contribution of the variant to the gene-based variant set signal. A quantitative linear model of mtDNA-CN was also fit for each study-wide significant gene, testing the conditional independence of each of variants with MAC ≥ 6 in a single model per gene and adjusting for age, sex, WES batch, and genotyping PCs 1-40.

### Gene set enrichment

We downloaded 33,750 gene sets from MitoCarta 3.0^4^, MsigDb v7.4^37^, and Disgenenet v7^38^. For each gene set, we compared means of z-scores from SMMAT tests for genes in and out of the set using a t-test assuming equal variance^39^. A single aggregate rare variant set, namely the CADD-PHRED ≥ 18 and MAF ≤ 0.001 variant set, was used to obtain z-scores. Since z-scores with extreme values can unduly influence the mean z-score of a gene set, we further tested for robustness of those resulting sets that were significantly enriched at a Bonferroni adjusted family-wise error rate (FWER) of 1.5 × 10^−6^ (85 gene sets) by excluding extreme-value SMMAT test z-scores (Supplemental Figure S3), specifically z-scores of *TFAM, JAK2, TWNK*, and *SAMHD1* tests, and re-computing a t-test enrichment p-value.

### PheWAS

The R package PheWAS^40^ was used to collapse summary diagnoses represented as distinct ICD10 codes in the UKB field 42170 into binary phecodes capturing cases and controls for a variety of disease categories. The analysis was restricted to unrelated individuals using the ‘used.in.pca.calculation’ variable^26^ in resource 531 of the UKB and was restricted to the White British subset of self-reported ancestry using UKB data field 21000. A logistic regression was run for each phecode as a dependent variable regressed onto the rare variant weighted sum score, adjusting for age, sex, and sequencing center. We used the simpleM^41^ method to compute the number of effective tests, which accounted for correlated phecode case and control statuses between clinically related and hierarchical phecodes. This yielded 1,530 effective tests which was used as the denominator in the Bonferroni multiple testing correction. The number of effective tests times the number of genes, i.e., p ≤ 3.0 × 10^−6^, was used as the Bonferroni multiple test correction denominator for the pheWAS of rare variant carrier status in individual genes. The R package MendelianRandomization^42^ was used to obtain the inverse variance weighted causal point estimate between mtDNA-CN rare variants in *SAMHD1* and the phecode for breast cancer risk.

All statistical analyses were performed in R 4.0.2.

## Results

### Sample characteristics

The current study included 415,422 self-reported White individuals (54.2% female) with a mean age of 56.8 (SD = 8.03) years enrolled in the UK Biobank (UKB)^26^. mtDNA-CN was estimated from DNA derived from blood buffy coat^12^.

### Single variants significantly associated with mtDNA-CN

We performed linear mixed-model association testing^29^ of single nucleotide variants (SNVs) and small insertion/deletions (indels) with minor allele count (MAC) ≥ 6 in 414,665 UK Biobank exomes of self-reported Whites, adjusting for fixed effects of age, sex, sequencing batch, and random effects due to genetic relatedness and ancestry. We also included genotypes at 129 previously discovered autosomal common variant GWAS loci^12^ as fixed effects to condition the results on independent genome-wide signals from common single variants associated with mtDNA-CN. We identified 11 SNVs associated with mtDNA-CN at a Bonferroni-corrected genome-wide significance (p ≤ 7.4 × 10^−9^) (Table 1A). The most significant SNVs were a pair of rare variants separated by 1.2Mb in *SAMHD1* (NC_000020.11:g.36893060C>T, p.Asp585Asn, beta = +0.61 SD [SE = 0.06], p-value = 4.2 × 10^−28^, MAC = 304) and *SPAG4* (NC_000020.11:g.35619007C>G, beta = +0.55 SD [SE = 0.06], p-value = 6.4 × 10^−22^, MAC = 288) across chromosome 20q11.22-q11.23 (Figure 1A).

**Table 1A.** Single nucleotide variants associated with mtDNA-CN at exome-wide significance (p ≤ 7.4 × 10^−9^).

|  | Chromosome | Position (bp)<sup>a</sup> | Reference Allele | Effect Allele | Effect Size<sup>b</sup> | Standard Error<sup>b</sup> | Minor Allele Count | P-value | Q-value | Locus / Gene | Functional Effect | Proportion Variance Explained<sup>c</sup> |
| --- | --- | --- | --- | --- | --- | --- | --- | --- | --- | --- | --- | --- |
| | 20 | 36,893,060<sup>d</sup> | C | T | 0.61 | 0.06 | 304 | $4.20 \times 10^{-28}$ | $2.84 \times 10^{-21}$ | *SAMHD1* | p.Asp585Asn | 0.029% |
| | 20 | 1,895,355 | G | C | -0.17 | 0.02 | 3,898 | $6.60 \times 10^{-24}$ | $2.23 \times 10^{-17}$ | *SIRPA* | Intronic variant | 0.024% |
| | 20 | 35,619,007<sup>d</sup> | C | G | 0.55 | 0.06 | 288 | $6.40 \times 10^{-22}$ | $1.44 \times 10^{-15}$ | *SPAG4* | Intronic variant | NA<sup>d</sup> |
| | 9 | 5,073,770 | G | T | 0.47 | 0.06 | 284 | $2.70 \times 10^{-17}$ | $4.57 \times 10^{-11}$ | *JAK2* | p.Val617Ile | 0.017% |
| | 10 | 100,993,257 | G | A | -1.07 | 0.14 | 48 | $1.50 \times 10^{-14}$ | $2.03 \times 10^{-8}$ | *TWNK* | p.Arg601Gln | 0.014% |
| | 17 | 18,291,048 | T | C | 0.06 | 0.01 | 17,340 | $6.40 \times 10^{-14}$ | $7.22 \times 10^{-8}$ | *TOP3A* | Intronic variant | 0.014% |
| | 4 | 184,766,776 | G | A | 2.36 | 0.37 | 7 | $7.80 \times 10^{-11}$ | $7.54 \times 10^{-5}$ | *ACSL1* | Intronic variant | 0.010% |
| | 20 | 36,898,455 | C | G | 0.25 | 0.04 | 599 | $3.50 \times 10^{-10}$ | $2.96 \times 10^{-4}$ | *SAMHD1* | p.Arg531Ser | 0.009% |
| | 20 | 34,280,961 | C | T | 2.45 | 0.4 | 6 | $5.60 \times 10^{-10}$ | $4.21 \times 10^{-4}$ | *AHCY* | 3' UTR variant | 0.009% |
| | 1 | 205,848,004 | A | C | 2.36 | 0.4 | 6 | $2.50 \times 10^{-9}$ | $1.69 \times 10^{-3}$ | *PM20D1* | Intronic variant | 0.009% |
| | 5 | 149,332,902 | C | A | 2.33 | 0.4 | 6 | $4.00 \times 10^{-9}$ | $2.46 \times 10^{-3}$ | *AFAP1L1* | Intronic variant | 0.008% |
| Total | — | — | — | — | — | — | — | — | — | — | — | 0.144% |

<sup>a</sup> Genome coordinates reported in GRCh38
<sup>b</sup> Effect sizes and standard errors are in units of standard deviation of the scaled mtDNA-CN distribution (mean = 0, std. dev. = 1)
<sup>c</sup> Proportion of variance (PVE) in mtDNA-CN explained by variant, computed as $PVE = \text{Effect-Size}^2 / (\text{Effect-Size}^2 + \text{Std.-Error}^2 * \text{Sample-Size})$ (n=414665)
<sup>d</sup> Variants present in a shared set of 262 individuals, as part of a rare, shared ancestral haplotype. *SPAG4* in linkage disequilibrium with *SAMHD1*.

**Figure 1.**
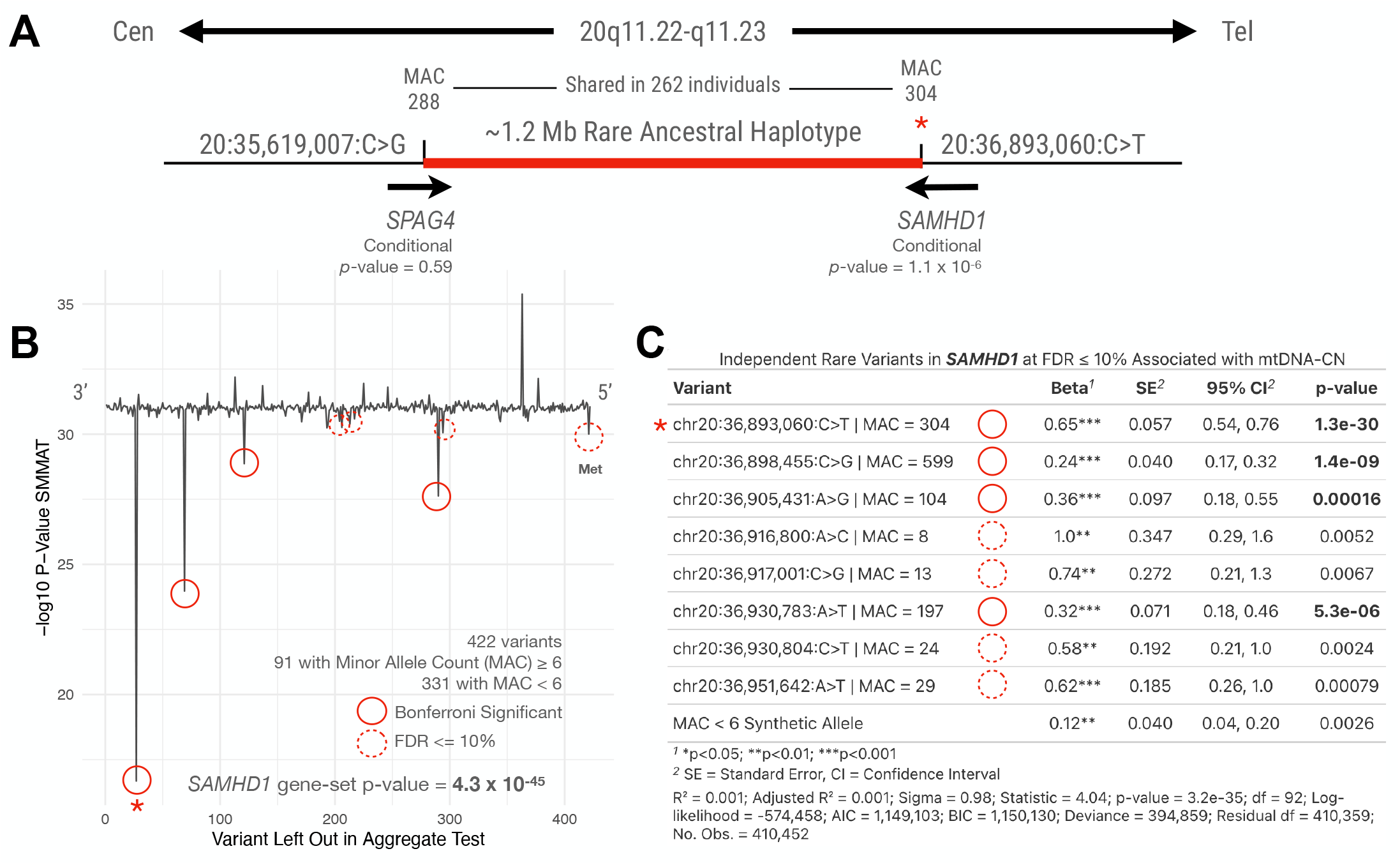
Rare, 1.2 Mb shared ancestral haplotype associated with increased mtDNA-CN. A) Rare variants in *SPAG4* (MAC = 288, association with mtDNA-CN p = 6.4 × 10^−22^, conditional p = 0.59) and *SAMHD1* (MAC = 304, association with mtDNA-CN p = 4.2 × 10^−28^, conditional p = 1.1 × 10^−6^) were present in a shared set of 262 individuals, representing an ancestral haplotype associated with mtDNA-CN. B) Fine mapping of gene-level rare variant effects via leave-one-out analysis of all 422 protein-coding nonsynonymous variants in *SAMHD1* identifies multiple independent variants associated with mtDNA-CN. Unbroken and dashed red circles indicate gene-level Bonferroni-significant and FDR ≤ 10% associations with mtDNA-CN, respectively. C) Conditional analysis of all 91 MAC ≥ 6 alleles and a MAC < 6 synthetic allele within a single linear model of mtDNA-CN adjusting for age, sex, ancestry PCs 1-40 and exome sequencing batch. Bold p-values highlight four Bonferroni-significant conditionally independent associations, including the missense variant on the rare, shared haplotype (red star). MAC = minor allele count, FDR = false discovery rate, Met = Methionine start codon, Cen = centromere, Tel = telomere.

Conditioning on the common variant signal allowed us to uncover an independent rare non-coding variant within intron one of *SIRPA* (NC_000020.11:g.1895355G>C, beta = -0.17 SD [SE = 0.02], p-value = 6.6 × 10^−24^, MAC = 3898), a locus previously identified through the common-variant GWAS^12^. Additional exome-wide significant single variant associations for mtDNA-CN included a missense variant in *JAK2* (NC_000009.12:g.5073770G>T, p.Val617Ile, beta = +0.47 SD [SE = 0.06], p-value = 2.7 × 10^−17^, MAC = 284), a known dominant somatic gain-of-function mutation associated with myeloproliferative disorders^43^, and a missense variant within the mtDNA helicase *TWNK* (NC_000010.11:g.100993257G>A, p.Arg601Gln, beta = -1.07 SD [SE = 0.14], p-value = 1.5 × 10^−14^, MAC = 48) associated with mitochondrial DNA depletion syndrome 7 (MTDPS7 [MIM: 271245]).

Four variants at study-wide significance were ultra-rare with MAC between six and seven and with concomitantly large effect sizes (mean beta = +2.4 SD, mean SE 0.4 SD). Given the low MAC of these variants, we tested for robustness of these findings by considering sources of false positive effects due to the influence of an outlier mtDNA-CN value, population substructure, or cryptic relatedness. All four variants were robust to the above sources of confounding (Supplemental Figure S4, Supplemental Table S2), and test statistics of variants at MAC 6, 7, and 8 each showed no signs of test statistic inflation (Supplemental Figures S5-6).

### Rare, ancestral haplotype on 20q11 significantly associated with mtDNA-CN

Given that the rare single variant associations in *SAMHD1* and *SPAG4* are in close physical proximity and have similar MAC, we investigated the independence of the variant effects on mtDNA-CN. The missense variants in *SPAG4* (MAC 288, p = 6.4 × 10^−22^) and *SAMHD1* (MAC 304, p = 4.2 × 10^−28^) were present in a shared set of 262 individuals (Figure 1A). The association with mtDNA-CN, however, was only significant among carriers of both variants (p = 9.94 × 10^−21^) and those with only the *SAMHD1* variant (40 heterozygotes, p = 0.002) (Supplemental Figure S7). Conditional analysis including both SNPs in the same model confirmed that the association to mtDNA-CN was driven by the *SAMHD1* variant (conditional p-value = 1.1 × 10^−6^, *SPAG4* conditional p-value = 0.59).

These data are consistent with the presence of a rare ancestral haplotype segregating in this population between *SPAG4* and *SAMHD1* that is associated with mtDNA-CN, with the association with mtDNA-CN at *SPAG4* solely due to its linkage disequilibrium with *SAMHD1*. Indeed, the 1.2 Mb region between *SPAG4* and *SAMHD1* lies in a previously described recombination desert (Supplemental Figure S8)^44^, and the surrounding DNA on 20q11 is therefore identical-by-descent in the subset of UK Biobank participants that share this haplotype.

### Gene-based groups of rare variants significantly associated with mtDNA-CN

We next implemented a linear mixed-model aggregate rare variant association testing framework^31^ to test for association between sets of rare variants in protein-coding genes and mtDNA-CN. A null mixed-effects model was fit accounting for random effects of genetic kinship and fixed effects of age, sex, sequencing batch, and genotype-derived principal components of ancestry. Due to a lack of priors on true causality that can be used to identify the ideal subset of rare variants to be included in a gene-based test, for each gene we tested 9 sets of nonsynonymous protein coding sequence variant combinations at increasing mutational severity thresholds (all nonsynonymous variants, nonsynonymous variants with CADD-PHRED score ≥ 18, predicted loss-of-function (pLoF) variants), and decreasing MAF (≤1%, ≤0.1%, ≤0.01%). Eight genes were associated with mtDNA-CN at a study-wide significance cutoff of p ≤ 6.7 × 10^−7^ (Table 1B). Three additional genes (*POLRMT*, p-value 1.1 × 10^−6^, *CAVIN2*, p-value 1.9 × 10^−6^, and *MGME1*, p-value 2.0 × 10^−6^) had marginal significance (Supplemental Table S3), of which two out of three (*POLRMT, MGME1*) have previously been linked to mitochondrial function. *POLRMT* encodes the RNA polymerase for mtDNA and is part of the mitochondrial transcription initiation complex which includes *TFAM* and *TFB2M* that is required for basal transcription of mitochondrial DNA^45^. *MGME1* is a mtDNA maintenance exonuclease and loss-of-function mutations within *MGME1* have been linked with autosomal recessive mtDNA depletion syndrome-11 (MTDPS11 [MIM: 615084])^46^.

**Table 1B.**
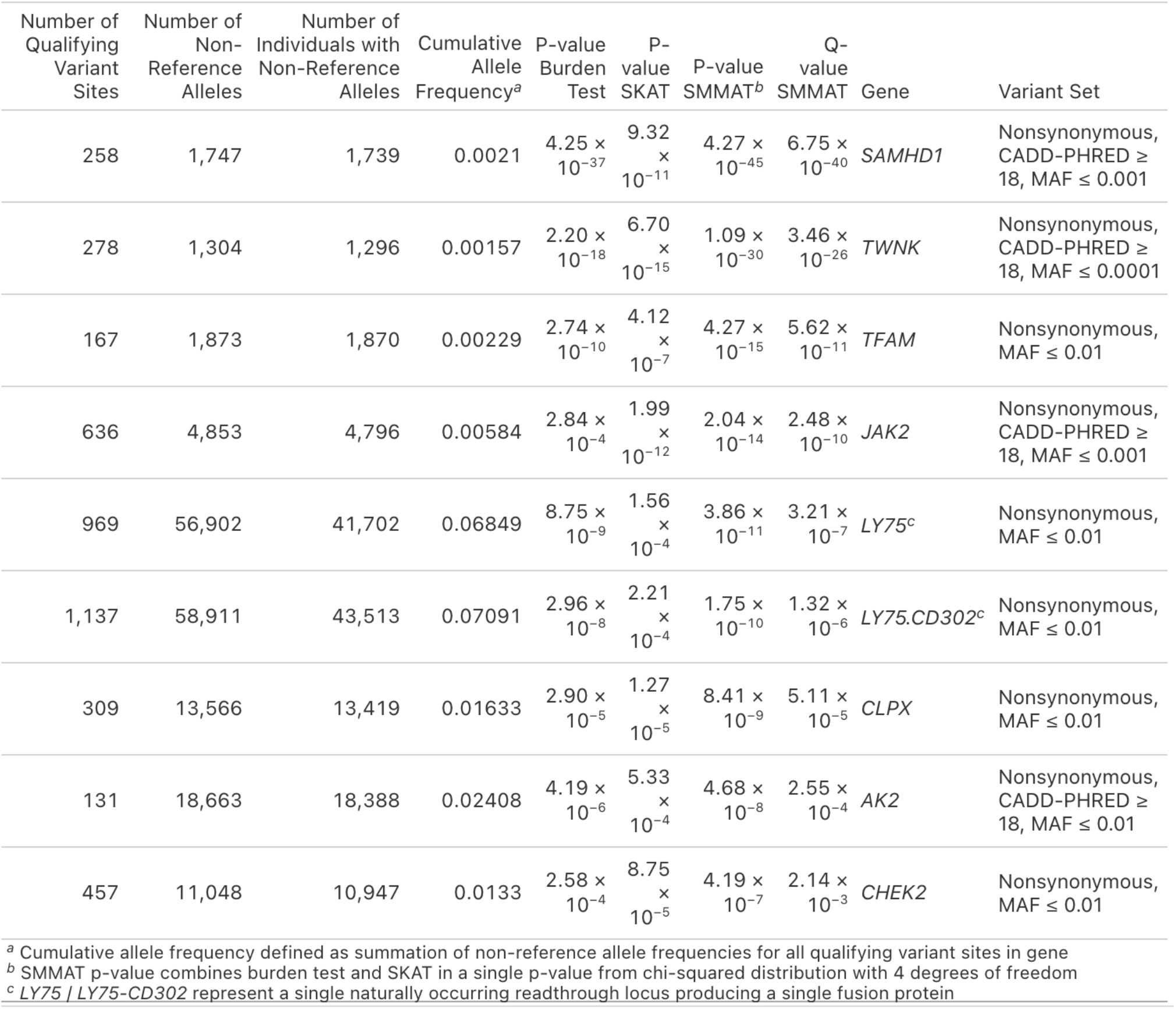
Aggregate gene-based rare variants associated with mtDNA-CN at study-wide significance (p ≤ 6.7 × 10^−7^).

For each gene, we used an iterative leave-one-out analysis to test the hypothesis that a subset of variants at each locus is gene-level conditionally independent and drives most of the association signal. Across eight genes, we identified 18 gene-level Bonferroni-significant conditionally independent associations (Supplemental Table S4), including 4 missense mutations in the most significantly associated gene, *SAMHD1* (gene-based test p-value = 4.3 × 10^−45^, Figures 1B-C). Notably, conditionally independent variants had consistent effect directions within a gene, in line with the recent observation of long allelic series of rare coding variants in a large-scale exome study^47^ in the UKB, and suggesting that independent alterations to distinct protein residues within the same gene affected mtDNA-CN in a consistent manner. Three genes (*CHEK2, POLRMT, CAVIN2*) had no individual variant that was significantly associated after gene-wide multiple-testing correction, suggesting a multitude of small-effects across the genes drove the gene set signal at these loci.

Most loci with conditionally independent effects had trait-increasing alleles, with only *TWNK* (SMMAT test p-value = 1.1 × 10^−30^) and *TFAM* (SMMAT test p-value = 4.3 × 10^−15^) harboring evidence of multiple independent mtDNA-CN lowering effect alleles, consistent with their role and localization in the mtDNA nucleoid and their implication in autosomal recessive Mendelian mtDNA depletion syndromes. Notably, only *TWNK* and *TFAM* harbored ultra-rare (MAC < 6) nonsynonymous variants that had significant association with mtDNA-CN. These MAC < 6 variants, coded as a single synthetic allele, were associated with lower mtDNA-CN in *TWNK* (beta = -0.25 SD [SE = 0.04], p-value = 2.17 × 10^−10^) and *TFAM* (beta = -0.37 SD [SE = 0.06], p-value = 8.3 × 10^−9^).

### Rare variants for mtDNA-CN significantly associated with Mendelian mtDNA depletion syndromes

To test the hypothesis that mtDNA-CN-associated rare variants are enriched in known causal mtDNA depletion syndrome genes, we tested whether the 20 known causal depletion syndrome genes^48^ have excess association signal in our data. Eight out of 20 genes were nominally associated at a p ≤ 0.05 (3 genes at a Bonferroni-adjusted p-value ≤ 0.05 accounting for 20 tests), consistent with a highly significant enrichment (exact binomial test p-value 2.9 × 10^−6^, Supplemental Table S5).

### Gene set enrichment analysis

We next tested for enrichment of aggregate gene-based rare variant signal within each of 33,750 annotated gene sets compiled from MsigDb^37^, Mito Carta^4^, and Disgenenet^38^ under the hypothesis that loci harboring rare variants for mtDNA-CN would implicate functionally relevant gene sets related to the genetic architecture of mtDNA-CN. Specifically, for each gene set, we compared means of z-scores from SMMAT tests for genes in and out of the set using a t-test assuming equal variance^39^. 85 gene sets were significantly associated at a Bonferroni adjusted family-wise error rate of 1.5 × 10^−6^ (Supplemental Table S6). To avoid implicating gene sets driven solely by a single gene with a large z-score, we re-computed the enrichment p-value for the 85 gene sets after excluding extreme value SMMAT test z-scores in each gene set (*TFAM, JAK2, TFAM*, and *SAMHD1*; Supplemental Figure S3). 19 gene sets retained significance after multiple test correction (p ≤ 0.05/85), including four that retained Bonferroni-significance accounting for 33,750 tests. Terms central to the synthesis and maintenance of mtDNA were the most significantly enriched, including mtDNA replication (p-value 1.6 × 10^−26^). mtDNA-CN associated genes were also enriched for medically relevant phenotypes including those related to mitochondrial disease, such as quadriceps muscle weakness (p-value 3.4 × 10^−21^) and progressive external ophthalmoplegia (p-value 2.5 × 10^−16^), and bleeding disorders such as macrothrombocytopenia (p-value 7.4 × 10^−7^). Finally, a gene set composed of common variant implicated loci^12^ was also significantly enriched for rare variant signal (n=111 genes in gene set, mean z-score of genes in set = 1.30, background z-score = 0.83, p-value = 3.04 × 10^−13^, p-value after removing extreme values = 1.02 × 10^−5^).

### Disease PheWAS

To test the hypothesis that rare genetic variation associated with mtDNA-CN highlights causal pathways between mtDNA-CN and disease risk, we collapsed summary diagnoses from inpatient hospital records represented by distinct ICD10 codes for 314,201 unrelated individuals of White British ancestry in the UKB into 1,623 broad phecodes. We then tested for association in a phenome-wide association study (pheWAS) between the phecodes and rare variant carrier status represented as a mtDNA-CN-associated rare variant summed score weighted by effect sizes, reasoning that rare variants affecting mtDNA-CN act additively to modulate risk of a common downstream phenotype. The weighted sum score was restricted to variants of an effect size larger than +/- 0.3 SD on mtDNA-CN and included a synthetic allele of ultra-rare variants with MAC < 6 in *TFAM* that had an effect size < -0.3 SD. PheWAS of the mtDNA-CN rare variant score yielded Bonferroni-significant associations to myeloproliferative disease (phecode 200) and myeloid leukemia (phecode 204.2). To assess whether these phecode associations were driven by rare variants at a single gene, we removed the variant at the *JAK2* locus which was individually strongly associated with the same phecodes (MAC = 284, OR = 822.4 [95% CI = 821.0 – 823.9], t-statistic = 36.9, p-value = 3.2 × 10^−299^ for phecode 200, and OR = 166.4 [95% CI = 164.6 – 168.2], t-statistic = 17.1, p-value = 8.5 × 10^−66^ for phecode 204.2) and noted that there were no significant associations to any disease phecodes after removing the *JAK2* variant from the weighted sum score, indicating that the association of the rare variant score to myeloproliferative disease was driven by the rare variant at the *JAK2* locus. We further ran pheWAS separately for each gene implicated by rare variation and found that carrier status of conditionally independent mtDNA-CN associated *SAMHD1* rare variants (MAC ≥ 6) was marginally associated with increased risk of breast cancer (phecode 174, k= 15,958 cases, OR = 2.1 [95% CI = 1.5 - 2.9], p-value 1.9 × 10^−5^). To assess whether any single conditionally independent variant at the *SAMHD1* locus drove this association, we tested each variant separately for breast cancer risk. Notably, there was a linear dose-response relationship with breast cancer risk for the four conditionally independent, uncorrelated mtDNA-CN associated variants at the *SAMHD1* locus (Figure 2, inverse-variance weighted Mendelian randomization causal point estimate = 1.29 [SE = 0.253], p=3.1 × 10^−7^, Supplemental Table S7).

**Figure 2.**
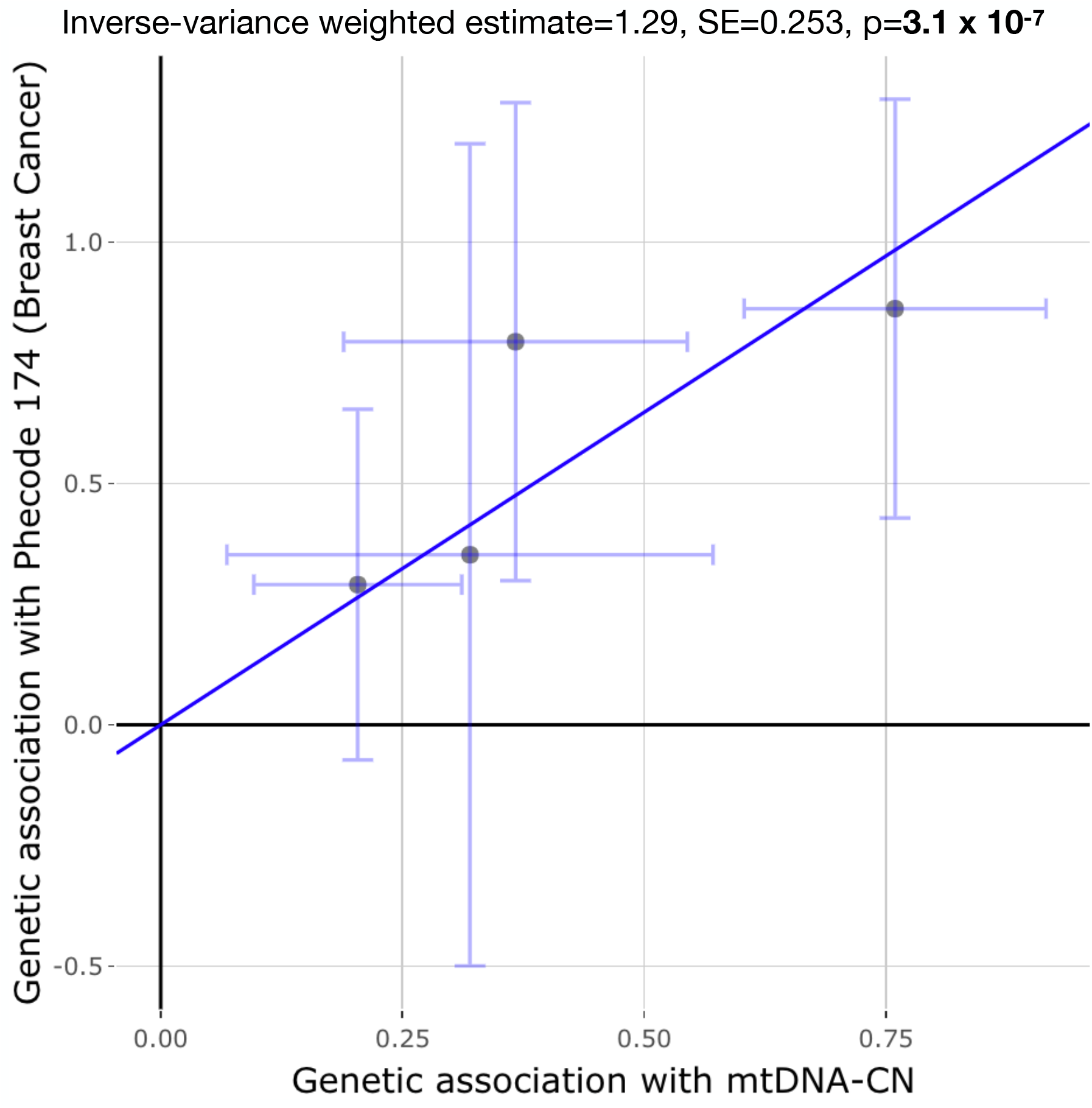
mtDNA-CN associated rare variants in *SAMHD1* exhibit a linear dose-response relationship with breast cancer risk. Effect estimates for gene-level conditionally independent associations with mtDNA-CN from four uncorrelated rare variants in *SAMHD1* are plotted against the corresponding association betas for disease phecode 174, representing breast cancer risk (n=225,098 females, k=17,517 cases; Supplemental Table S7). The slope of blue line illustrates the inverse-variance weighted causal estimate, bars around points represent 95% confidence intervals.

## Discussion

We conducted an exome-based study of genetic variation associated with mtDNA-CN in 415,422 individuals from the UK Biobank. We report 24 independent signals from 17 loci, including nine SNVs and one rare noncoding variant identified as study-wide independent. Using an iterative leave-one-out approach, we fine-mapped 14 gene-level conditionally independent effects. Rare variants were enriched in loci implicated in Mendelian mtDNA depletion syndromes, and these variants further implicated core processes involved with mtDNA replication, nucleoid structure formation, and maintenance. We note that while the manuscript was in preparation, Chong et al. 2022 identified *SAMHD1* from 147,740 samples as the only exome-based association signal for mtDNA-CN estimated using a different approach^49^, which provides additional evidence for our observed association of *SAMHD1* associated with mtDNA-CN. In part due to the ∼3x larger sample size, this study finds multiple additional loci. Combined with gene-based tests, we also identify 10 loci not detectable by common variant GWAS^12^ (MAF < 0.002), showcasing the added value of rare variants for discovery of loci contributing to the genetic architecture of mtDNA-CN.

In contrast to the expectation that physically distant rare variants are likely to be in linkage equilibrium^50,51^, we identified a pair of rare variants residing in a unique shared haplotype associated with mtDNA-CN. The variants in *SPAG4* and *SAMHD1* were separated by 1.2Mb on 20q11, and formed a single rare, shared haplotype in 262 individuals (haplotype frequency 3.2 × 10^−4^). The presence of a shared ancestral haplotype is supported by a similar MAC pattern for the rare variants in *SPAG4* and *SAMHD1* within the publicly available TOPMed Freeze 8 data (MAC = 21 and MAC = 25 for NC_000020.11:g.35619007C>G and NC_000020.11:g.36893060C>T, respectively), suggesting that this haplotype is older than 5-10 generations since the TOPMed cohort is >80% US residents who experienced a bottleneck during the migration to America^52^. Indeed, the shared haplotype lies in a known region of very low recombination on 20q^44^, suggesting the persistence of the identical-by-descent segment in this population bearing this haplotype can be attributed to the low recombination rate. This finding warrants further inspection of rare haplotype sharing affecting complex traits in large biobank-scale sequencing datasets.

The association with mtDNA-CN by the rare haplotype was solely driven by the *SAMHD1* mutation (NP_056289.2:p.Asp585Asn), which offers insight into the molecular mechanism behind how variation in *SAMHD1* leads to increased mtDNA-CN. *SAMHD1* encodes a deoxynucleoside triphosphate triphosphohydrolase, a host restriction factor enzyme that depletes dNTP pools and aids in the viral defense response^53^. The deoxynucleoside products of the enzymatic hydrolysis by SAMHD1 are transported into the mitochondria and salvaged to dNTPs^54^. While an overabundance of dNTPs could indicate a putative pathway by which heterozygous mutations within *SAMHD1* lead to increased mtDNA-CN, since one determinant of mtDNA-CN is nucleotide metabolism^12^, we further found that the p.Asp585Asn variant falls within a region of *SAMHD1* deleted in autosomal recessive Aicardi-Goutières syndrome 5 (AGS5 [MIM: 612952]). In particular, a homozygous p.Gln548Ter variant in the same region of the gene found in a *SAMHD1*^-/-^ AGS5 patient was demonstrated to influence SAMHD1’s alternative role in degrading nascent single-stranded DNA (ssDNA) at stalled replication forks but not influence dNTPase activity^55^, suggesting that the conditionally independent variant in our study may have an alternative molecular mechanism revolving around failure of ssDNA degradation.

*SAMHD1*’s role as a regulator of the innate immune response and a key part of the replication stress response also supports its role in cancer^56–58^ and allows for the consideration of the hypothesis that mtDNA-CN may be causally associated with cancer. Notably, conditional analysis of rare variants within *SAMHD1* revealed independent mtDNA-CN-increasing effects at four sites within *SAMHD1*, and there was a linear dose-response relationship between the four variants’ effects on mtDNA-CN and on breast cancer risk. These data support a mechanistic link between genetic variation at the *SAMHD1* locus on mtDNA-CN and on breast cancer. Importantly, however, we did not find a disease pheWAS signal for breast cancer among individuals carrying non-*SAMHD1* mtDNA-CN associated rare variants, suggesting mtDNA-CN is not causal for breast cancer risk, but that the mechanism that impacts mtDNA-CN due to SAMHD1 deficiency is having a proportional impact on breast cancer risk at the *SAMHD1* locus.

We also found a subset of other genes harboring mtDNA-CN associated rare variants in our study (*JAK2, TOP3A, CHEK2, SIRPA*) that are pleiotropic for cancer phenotypes. The missense mutation at the *JAK2* locus, p.Val617Ile, is a dominant gain-of-function mutation that is causal for myeloproliferative disease^43^, possibly a result of clonal hematopoeisis of intermediate potential (CHIP)^59,60^. Biallelic mutations at *TOP3A* have been associated with a Bloom-syndrome-like disorder^61^, a rare disorder characterized by predisposition to cancer, albeit with additional mitochondrial dysfunction consistent with the TOP3A function in mtDNA decatenation^62^. *CHEK2* is a tumor suppressor gene that encodes a checkpoint protein kinase activated due to DNA damage and controls cell cycle arrest, and mutations in *CHEK2* have been associated with autosomal dominant forms of breast, colorectal, and prostate cancer as part of familial Li-Fraumeni syndrome (LFS, [MIM: 151623])^63^. And *SIRPA*, a gene previously identified in the common variant GWAS for mtDNA-CN^12^, encodes a cell surface receptor for CD47 involved in prevention of auto-phagocytosis of myeloid cells that is frequently downregulated in a variety of cancers^64^. Nevertheless, while we found individual pleiotropic associations with cancer at loci implicated by rare variant associations with mtDNA-CN, combined, the mtDNA-CN rare variant weighted sum score was not associated with any cancer phecodes, lending credence to a competing hypothesis that mtDNA-CN is significantly dysregulated in a variety of cancers and that it is downstream of pathways leading to cancer.

Finally, studying rare variants associated with mtDNA-CN allowed us to pinpoint functional variants in known genes implicated in autosomal recessive Mendelian mtDNA depletion syndromes (*TWNK, TFAM*). The most significant *TWNK* variant decreasing mtDNA-CN was a missense mutation at NC_000010.11:g.100993257G>A (MAC = 48, beta = -1.09 SD [SE = 0.14], p-value = 1.4 × 10^−14^), a variant previously reported in two affected siblings with a diagnosis of autosomal recessive Perrault syndrome-5^65^ (PRLTS5, [MIM: 616138]) who each harbored an additional compound heterozygous mutation in *TWNK* and whose clinical presentation included muscle weakness and atrophy reminiscent of the broader mitochondrial DNA depletion syndrome-7 (MTDPS7, [MIM: 271245]), the autosomal recessive disorder caused by mutations in *TWNK. TFAM* encodes a transcription factor part of the initiation complex required for basal transcription of mitochondrial DNA and is a key component of mtDNA nucleoids^66,67^, and mutations within *TFAM* have previously been linked with mtDNA depletion syndrome-15 (MTDPS15, [MIM: 617156])^68,69^. Two variants were gene-level conditionally independent at *TFAM*, namely at NC_000010.11:g.58388676C>G (MAC = 320, aa residue Q100X, beta = -0.27 SD [SE = 0.058], p-value = 3.0 × 10^−6^) and at NC_000010.11:g.58394946A>G (MAC = 81, aa residue K173E, beta = -0.49 SD [SE=0.11), p-value = 1.3 × 10^−5^) that each fall within the experimentally validated DNA-binding domains^66,68^ for *TFAM*: the high mobility group (HMG) box 1 (residues 50-118) and HMG box 2 (residues 155-219), respectively. The conditional independence of the decreasing mtDNA-CN effects from variants in each of the DNA binding domains within TFAM confirms both residues are critical for proper functioning of the protein.

Several limitations exist for this study. First, the SNP-based heritability of the measure of mtDNA-CN used in this study is 7.0-7.4% as estimated by common variants^12^ which is in contrast to heritability estimates of 54-65% from previous family-based studies for mtDNA-CN^23,24^. Notwithstanding artificial inflation of heritability estimates from twin and family-based studies^70,71^, this gap suggests that, among other sources of genetic variation which could contribute to the missing heritability, including rare variants^25^, an imperfect phenotypic measurement affects the association test power of both common and rare variant GWAS. mtDNA-CN is ‘noisy’ due to confounding from platelet counts and other types of cells^27^, and while we account for these cell counts in our measurement, hidden sources of variability were not considered which might impact association test power. Analogously, blood mtDNA-CN has other determinants beyond mitochondrial function^27^ which limits interpretation of the rare genetic variants impacting the phenotype.

Second, despite the large sample size of the study, the power of the study to detect rare variant associations for mtDNA-CN is still limited. We investigated this deficit by extrapolating a power-law fit from the effect sizes and MAF of the 129 autosomal common variants for mtDNA-CN and estimated power along the spectrum of MAF and effect sizes expected for a presumed rare variant genetic architecture. For a study cohort size of 414,665 individuals and controlling type-I error at a Bonferroni-corrected alpha of 7.4 × 10^−9^, we noted that the present study is underpowered to detect rare single variant associations with mtDNA-CN along the predicted effect sizes for rare variants with MAF < 1% (Supplemental Figure S1). Finally, the study is limited to self-reported White individuals due to a lack of sample size for other genetic ancestry groups.

In conclusion, we have conducted the largest-to-date study of rare variation impacting mtDNA-CN, surveying over 400,000 individuals sourced from a single tissue and collection method. Using an association testing framework that tested single and gene-based groups of variants, seventeen genes were identified. These genes highlight the biological underpinnings of mtDNA-CN and mitochondrial function, including enrichment in genes associated with Mendelian mtDNA depletion syndrome genes. Combined, these data highlight the importance of studying rare genetic variation in the study of the complex mtDNA-CN phenotype.

## Data Availability

Whole exome sequencing data from the UK Biobank (UKB) are available for access through the UKB Research Analysis Platform with a completed UKB application. Summary statistics for mtDNA-CN rare variant association tests are available for download at https://www.arkinglab.org/resources/. Code, including scripts for generation of association statistics and additional statistical analyses, is available at https://github.com/ArkingLab/mtDNA-CN-ExWAS.

https://github.com/ArkingLab/mtDNA-CN-ExWAS

## Data and code availability

Whole exome sequencing data for the UKB is available for access through the UKB Research Analysis Platform with an approved UKB application. Summary statistics for mtDNA-CN rare variant association tests are available for download at https://www.arkinglab.org/resources/. Scripts and code for generation of association statistics and additional statistical analyses will be available at https://github.com/ArkingLab/mtDNA-CN-ExWAS.

## Author contributions

D.E.A. and V.P. conceptualized the study, designed the experiments, and edited the manuscript. V.P. conducted the experiments, performed formal statistical analyses, including design, creation and execution of all scripts and code, and wrote and edited the manuscript. W.S., C.S., S.Y., J.L. conducted the experiments, curated data, and provided technical and statistical analysis support. D.E.A., E.G., and N.P. designed experiments, provided research supervision, and edited the manuscript. All authors reviewed the manuscript.

## Web resources

UKB Research Access Platform, https://www.ukbiobank.ac.uk/enable-your-research/research-analysis-platform

Mitocarta 3.0, https://www.broadinstitute.org/mitocarta/mitocarta30-inventory-mammalian-mitochondrial-proteins-and-pathways

MSigDB v7.4, https://www.gsea-msigdb.org/gsea/msigdb/

DisGeNET v7, https://www.disgenet.org

OMIM, https://omim.org

VEP, https://www.ensembl.org/info/docs/tools/vep/index.html

BOLT-LMM, https://alkesgroup.broadinstitute.org/BOLT-LMM/

GENESIS, https://bioconductor.org/packages/release/bioc/html/GENESIS.html

## REFERENCES

1. Wang, C. & Youle, R. J. The Role of Mitochondria in Apoptosis. Annu. Rev. Genet. 43, 95–118 (2009).

2. Fang, C., Wei, X. & Wei, Y. Mitochondrial DNA in the regulation of innate immune responses. Protein Cell 7, 11–16 (2016).

3. West, A. P. et al. Mitochondrial DNA stress primes the antiviral innate immune response. Nature 520, 553–557 (2015).

4. Rath, S. et al. MitoCarta3.0: an updated mitochondrial proteome now with sub-organelle localization and pathway annotations. Nucleic Acids Res. 49, D1541–D1547 (2020).

5. Allen, J. F. Why chloroplasts and mitochondria retain their own genomes and genetic systems: Colocation for redox regulation of gene expression. Proc. Natl. Acad. Sci. 112, 10231–10238 (2015).

6. Friedman, J. R. & Nunnari, J. Mitochondrial form and function. Nature 505, 335–343 (2014).

7. Yonova-Doing, E. et al. An atlas of mitochondrial DNA genotype-phenotype associations in the UK Biobank. Nat. Genet. 53, 982–993 (2021).

8. El-Hattab, A. W. & Scaglia, F. Mitochondrial DNA Depletion Syndromes: Review and Updates of Genetic Basis, Manifestations, and Therapeutic Options. Neurotherapeutics 10, 186–198 (2013).

9. López-Otín, C., Blasco, M. A., Partridge, L., Serrano, M. & Kroemer, G. The Hallmarks of Aging. Cell 153, 1194–1217 (2013).

10. Laricchia, K. M. et al. Mitochondrial DNA variation across 56,434 individuals in gnomAD. Genome Res. gr.276013.121 (2022) doi:10.1101/gr.276013.121.

11. Liu, X. et al. Association of mitochondrial DNA copy number with cardiometabolic diseases. Cell Genomics 1, 100006 (2021).

12. Longchamps, R. J. et al. Genome-wide analysis of mitochondrial DNA copy number reveals loci implicated in nucleotide metabolism, platelet activation, and megakaryocyte proliferation. Hum. Genet. 141, 127–146 (2022).

13. Hägg, S., Jylhävä, J., Wang, Y., Czene, K. & Grassmann, F. Deciphering the genetic and epidemiological landscape of mitochondrial DNA abundance. Hum. Genet. 140, 849–861 (2021).

14. Ashar, F. N. et al. Association of Mitochondrial DNA Copy Number With Cardiovascular Disease. JAMA Cardiol. 2, 1247–1255 (2017).

15. Hong, Y. S. et al. Mitochondrial DNA Copy Number and Incident Heart Failure: The Atherosclerosis Risk in Communities (ARIC) Study. Circulation 141, 1823–1825 (2020).

16. Zhao, D. et al. Mitochondrial DNA copy number and incident atrial fibrillation. BMC Med. 18, 246 (2020).

17. Ashar, F. N. et al. Association of Mitochondrial DNA levels with Frailty and All-Cause Mortality. J. Mol. Med. Berl. Ger. 93, 177–186 (2015).

18. Pyle, A. et al. Reduced mitochondrial DNA copy number is a biomarker of Parkinson’s disease. Neurobiol. Aging 38, 216.e7-216.e10 (2016).

19. Wei, W. et al. Mitochondrial DNA point mutations and relative copy number in 1363 disease and control human brains. Acta Neuropathol. Commun. 5, 13 (2017).

20. Yang, S. Y. et al. Blood-derived mitochondrial DNA copy number is associated with gene expression across multiple tissues and is predictive for incident neurodegenerative disease. Genome Res. 31, 349–358 (2021).

21. Reznik, E. et al. Mitochondrial DNA copy number variation across human cancers. eLife 5, e10769 (2016).

22. Hu, L., Yao, X. & Shen, Y. Altered mitochondrial DNA copy number contributes to human cancer risk: evidence from an updated meta-analysis. Sci. Rep. 6, 35859 (2016).

23. Ding, J. et al. Assessing Mitochondrial DNA Variation and Copy Number in Lymphocytes of ∼2,000 Sardinians Using Tailored Sequencing Analysis Tools. PLOS Genet. 11, e1005306 (2015).

24. Xing, J. et al. Mitochondrial DNA content: its genetic heritability and association with renal cell carcinoma. J. Natl. Cancer Inst. 100, 1104–1112 (2008).

25. Wainschtein, P. et al. Assessing the contribution of rare variants to complex trait heritability from whole-genome sequence data. Nat. Genet. 54, 263–273 (2022).

26. Bycroft, C. et al. The UK Biobank resource with deep phenotyping and genomic data. Nature 562, 203–209 (2018).

27. Picard, M. Blood mitochondrial DNA copy number: What are we counting? Mitochondrion 60, 1– 11 (2021).

28. Moore, C. M., Jacobson, S. A. & Fingerlin, T. E. Power and Sample Size Calculations for Genetic Association Studies in the Presence of Genetic Model Misspecification. Hum. Hered. 84, 256–271 (2019).

29. Loh, P.-R. et al. Efficient Bayesian mixed-model analysis increases association power in large cohorts. Nat. Genet. 47, 284–290 (2015).

30. Willer, C. J., Li, Y. & Abecasis, G. R. METAL: fast and efficient meta-analysis of genomewide association scans. Bioinformatics 26, 2190–2191 (2010).

31. Gogarten, S. M. et al. Genetic association testing using the GENESIS R/Bioconductor package. Bioinformatics 35, 5346–5348 (2019).

32. Frankish, A. et al. GENCODE 2021. Nucleic Acids Res. 49, D916–D923 (2021).

33. Lee, S., Abecasis, G. R., Boehnke, M. & Lin, X. Rare-Variant Association Analysis: Study Designs and Statistical Tests. Am. J. Hum. Genet. 95, 5–23 (2014).

34. Wu, M. C. et al. Rare-Variant Association Testing for Sequencing Data with the Sequence Kernel Association Test. Am. J. Hum. Genet. 89, 82–93 (2011).

35. Chen, H. et al. Efficient Variant Set Mixed Model Association Tests for Continuous and Binary Traits in Large-Scale Whole-Genome Sequencing Studies. Am. J. Hum. Genet. 104, 260–274 (2019).

36. Rentzsch, P., Witten, D., Cooper, G. M., Shendure, J. & Kircher, M. CADD: predicting the deleteriousness of variants throughout the human genome. Nucleic Acids Res. 47, D886–D894 (2019).

37. Liberzon, A. et al. Molecular signatures database (MSigDB) 3.0. Bioinformatics 27, 1739–1740 (2011).

38. Piñero, J. et al. The DisGeNET knowledge platform for disease genomics: 2019 update. Nucleic Acids Res. 48, D845–D855 (2020).

39. Irizarry, R. A., Wang, C., Zhou, Y. & Speed, T. P. Gene Set Enrichment Analysis Made Simple. Stat. Methods Med. Res. 18, 565–575 (2009).

40. Carroll, R. J., Bastarache, L. & Denny, J. C. R PheWAS: data analysis and plotting tools for phenome-wide association studies in the R environment. Bioinformatics 30, 2375–2376 (2014).

41. Gao, X., Starmer, J. & Martin, E. R. A multiple testing correction method for genetic association studies using correlated single nucleotide polymorphisms. Genet. Epidemiol. 32, 361–369 (2008).

42. Yavorska, O. O. & Burgess, S. MendelianRandomization: an R package for performing Mendelian randomization analyses using summarized data. Int. J. Epidemiol. 46, 1734–1739 (2017).

43. Kralovics, R. et al. A Gain-of-Function Mutation of JAK2 in Myeloproliferative Disorders. N. Engl. J. Med. 352, 1779–1790 (2005).

44. Deloukas, P. et al. The DNA sequence and comparative analysis of human chromosome 20. 414, 8 (2001).

45. Hillen, H. S., Morozov, Y. I., Sarfallah, A., Temiakov, D. & Cramer, P. Structural Basis of Mitochondrial Transcription Initiation. Cell 171, 1072-1081.e10 (2017).

46. Kornblum, C. et al. Loss-of-function mutations in MGME1 impair mtDNA replication and cause multisystemic mitochondrial disease. Nat. Genet. 45, 214–219 (2013).

47. Barton, A. R., Sherman, M. A., Mukamel, R. E. & Loh, P.-R. Whole-exome imputation within UK Biobank powers rare coding variant association and fine-mapping analyses. Nat. Genet. 53, 1260– 1269 (2021).

48. OMIM Phenotypic Series - PS603041. https://omim.org/phenotypicSeries/PS603041.

49. Chong, M. et al. GWAS and ExWAS of blood mitochondrial DNA copy number identifies 71 loci and highlights a potential causal role in dementia. eLife 11, e70382 (2022).

50. Li, B., Liu, D. J. & Leal, S. M. Identifying Rare Variants Associated with Complex Traits via Sequencing. Curr. Protoc. Hum. Genet. 78, 1.26.1-1.26.22 (2013).

51. Turkmen, A. & Lin, S. Are rare variants really independent? Genet. Epidemiol. 41, 363–371 (2017).

52. NHLBI Trans-Omics for Precision Medicine (TOPMed) Consortium et al. Sequencing of 53,831 diverse genomes from the NHLBI TOPMed Program. Nature 590, 290–299 (2021).

53. Franzolin, E. et al. The deoxynucleotide triphosphohydrolase SAMHD1 is a major regulator of DNA precursor pools in mammalian cells. Proc. Natl. Acad. Sci. 110, 14272–14277 (2013).

54. Franzolin, E., Salata, C., Bianchi, V. & Rampazzo, C. The Deoxynucleoside Triphosphate Triphosphohydrolase Activity of SAMHD1 Protein Contributes to the Mitochondrial DNA Depletion Associated with Genetic Deficiency of Deoxyguanosine Kinase. J. Biol. Chem. 290, 25986–25996 (2015).

55. Coquel, F. et al. SAMHD1 acts at stalled replication forks to prevent interferon induction. Nature 557, 57–61 (2018).

56. Clifford, R. et al. SAMHD1 is mutated recurrently in chronic lymphocytic leukemia and is involved in response to DNA damage. Blood 123, 1021–1031 (2014).

57. Rentoft, M. et al. Heterozygous colon cancer-associated mutations of SAMHD1 have functional significance. Proc. Natl. Acad. Sci. 113, 4723–4728 (2016).

58. Wang, J., Lu, F., Shen, X.-Y., Wu, Y. & Zhao, L. SAMHD1 is down regulated in lung cancer by methylation and inhibits tumor cell proliferation. Biochem. Biophys. Res. Commun. 455, 229–233 (2014).

59. Genovese, G. et al. Clonal Hematopoiesis and Blood-Cancer Risk Inferred from Blood DNA Sequence. N. Engl. J. Med. 371, 2477–2487 (2014).

60. Jaiswal, S. et al. Clonal hematopoiesis and risk for atherosclerotic cardiovascular disease. N. Engl. J. Med. 377, 111–121 (2017).

61. Martin, C.-A. et al. Mutations in TOP3A Cause a Bloom Syndrome-like Disorder. Am. J. Hum. Genet. 103, 221–231 (2018).

62. Nicholls, T. J. et al. Topoisomerase 3α Is Required for Decatenation and Segregation of Human mtDNA. Mol. Cell 69, 9-23.e6 (2018).

63. Bell, D. W. et al. Heterozygous germ line hCHK2 mutations in Li-Fraumeni syndrome. Science 286, 2528–2531 (1999).

64. Takahashi, S. Molecular functions of SIRPα and its role in cancer. Biomed. Rep. 9, 3–7 (2018).

65. Oldak, M. et al. Novel neuro-audiological findings and further evidence for TWNK involvement in Perrault syndrome. J. Transl. Med. 15, 25 (2017).

66. Vozáriková, V. et al. Mitochondrial HMG-Box Containing Proteins: From Biochemical Properties to the Roles in Human Diseases. Biomolecules 10, 1193 (2020).

67. Campbell, C. T., Kolesar, J. E. & Kaufman, B. A. Mitochondrial transcription factor A regulates mitochondrial transcription initiation, DNA packaging, and genome copy number. Biochim. Biophys. Acta BBA - Gene Regul. Mech. 1819, 921–929 (2012).

68. Larsson, N.-G. et al. Mitochondrial transcription factor A is necessary for mtDNA maintenance and embryogenesis in mice. 18, 6 (1998).

69. Stiles, A. R. et al. Mutations in TFAM, encoding mitochondrial transcription factor A, cause neonatal liver failure associated with mtDNA depletion. Mol. Genet. Metab. 119, 91–99 (2016).

70. Mayhew, A. J. & Meyre, D. Assessing the Heritability of Complex Traits in Humans: Methodological Challenges and Opportunities. Curr. Genomics 18, 332–340 (2017).

71. Manolio, T. A. et al. Finding the missing heritability of complex diseases. Nature 461, 747–753 (2009).

