## Supplementary Figures for "Whole-exome sequencing in 415,422 individuals identifies rare variants associated with mitochondrial DNA copy number"

**Figure S1. Study power as a function of minor allele frequency and absolute effect size.**

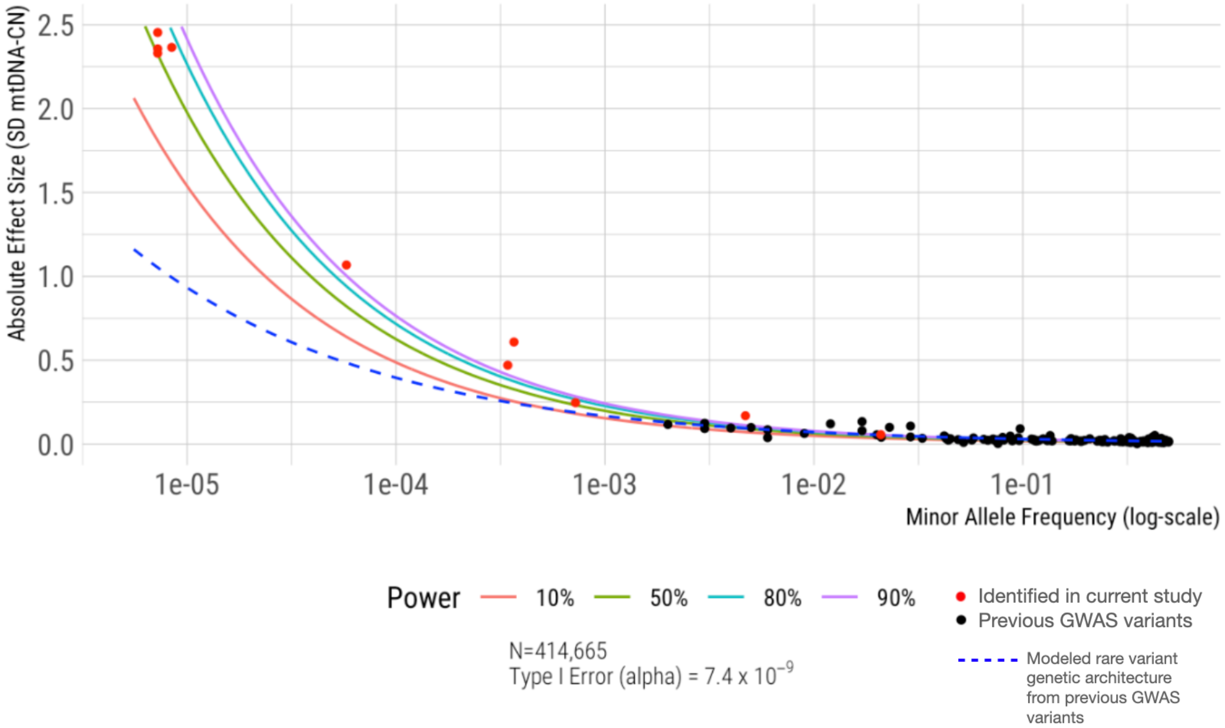


Statistical power isocurves are plotted at 10%, 50%, 80%, and 90% power for a study size of N=414,665 and controlling Type-I error at $7.4 \times 10^{-9}$ (Bonferroni-adjusted cutoff for 6,766,699 single variant tests) assuming a dominant model. Blue dashed line indicates power-law fit of 129 common autosomal variants previously independently associated with mtDNA-CN[1] (black points). Red points indicate single variant associations identified in the current study.

**Figure S2. Correlation between SMMAT test p-values across 9 variant sets.**

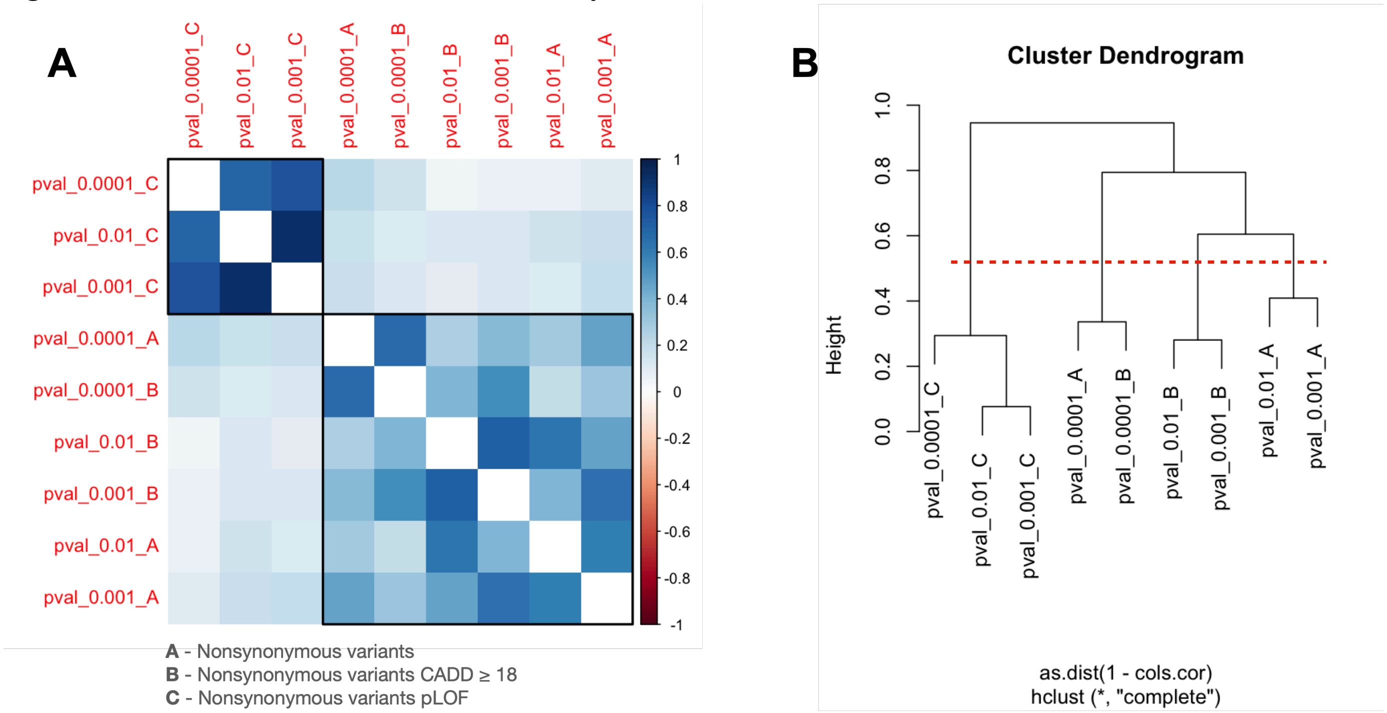


A) Correlation plot of p-values for 13,581 common genes from nine variant set combination tests chosen to cover "A" = nonsynonymous variants, "B" = nonsynonymous variants at CADD-PHRED ≥ 18, "C" = predicted loss-of-function (pLOF) nonsynonymous variants at MAF cutoffs of 0.01, 0.001, and 0.0001. Boxes represent two broad clusters as seen in B) Hierarchical clustering of dissimilarity computed as 1 – correlation between the nine variant-set p-values. Red dash line indicates chosen clustering height cutoff identifying four distinct clusters of p-values.

**Figure S3 Distribution of SMMAT Z-scores for gene-set enrichment analysis.**

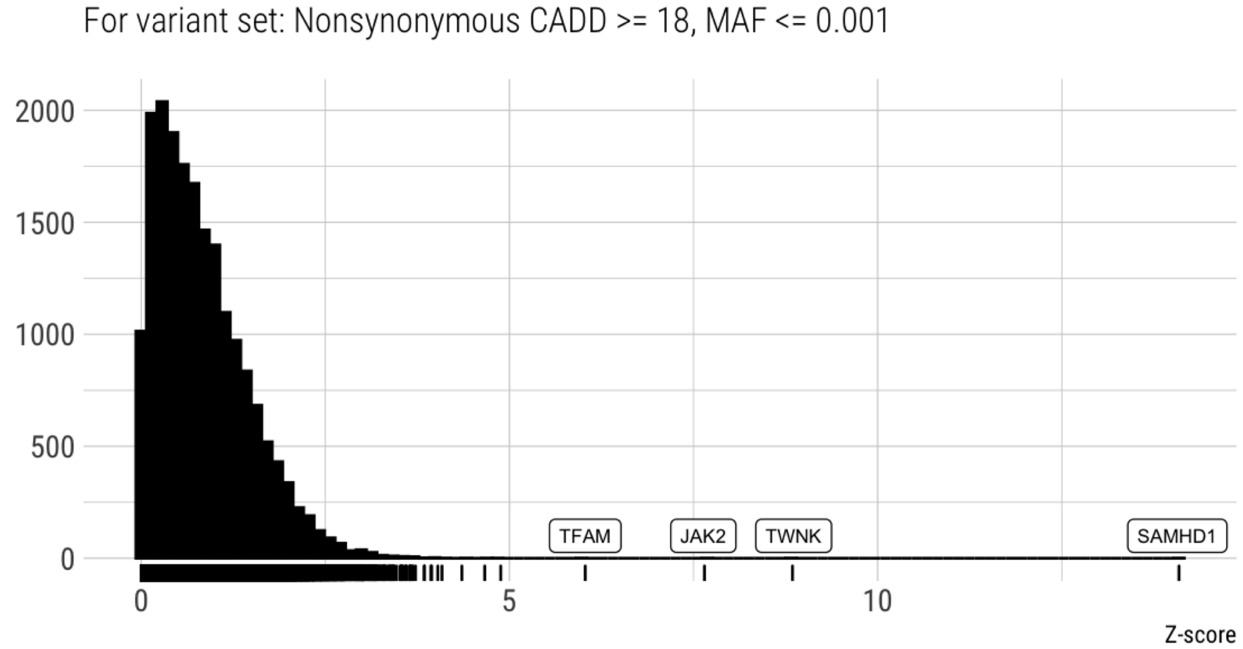


Z-score distribution demonstrates the presence of extreme values from the most significantly associated genes (*TFAM*, *JAK2*, *TWNK*, and *SAMHD1*).

**Figure S4. mtDNA-CN distributions for genome-wide significant ultra-rare MAC 6-7 variants.**

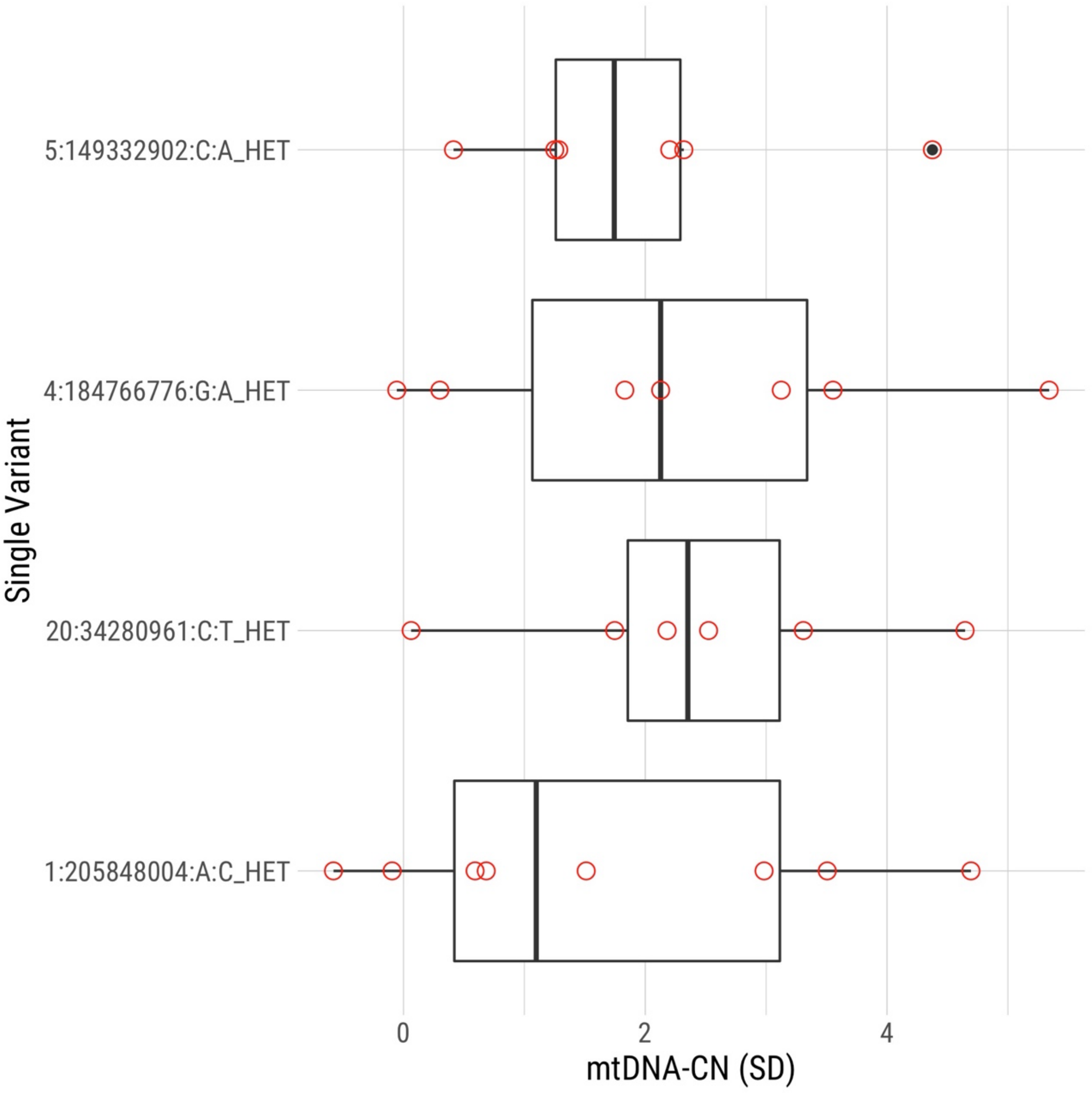

**Figure S5. QQ plot of rare variant associations stratified by minor allele count (MAC).**

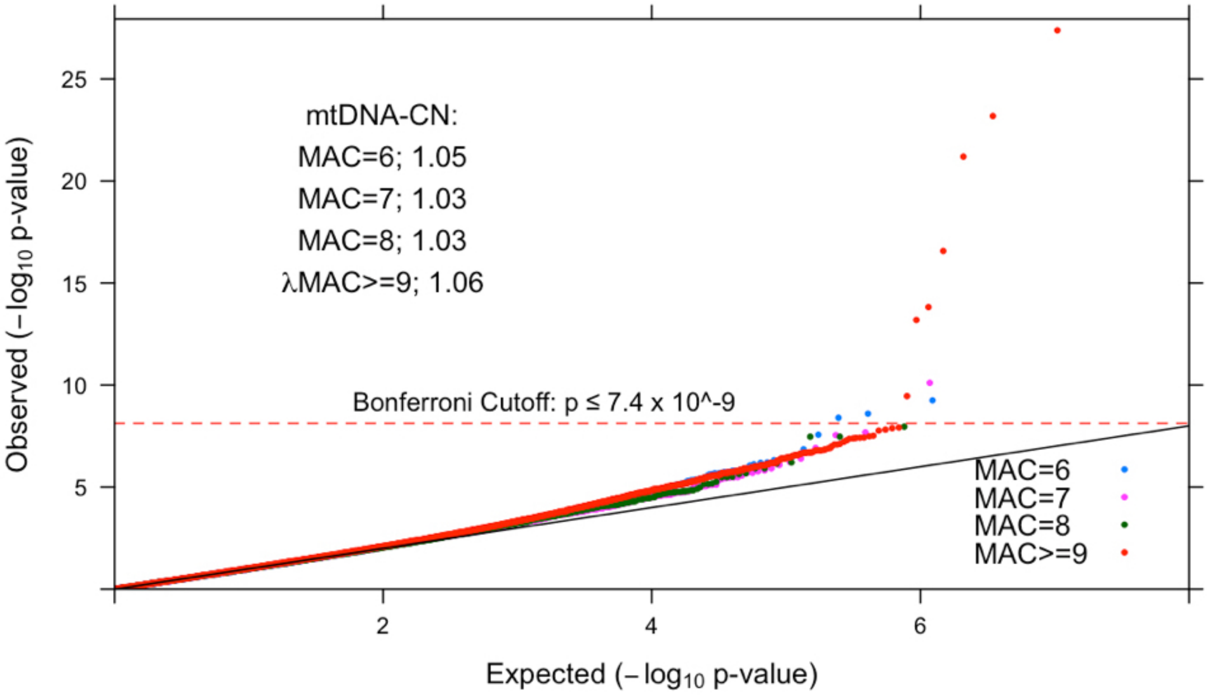

**Figure S6. QQ plot of rare variant associations with non-inverse normal transformed phenotype, inverse normal transformed mtDNA-CN phenotype, and in White British subset of individuals.**

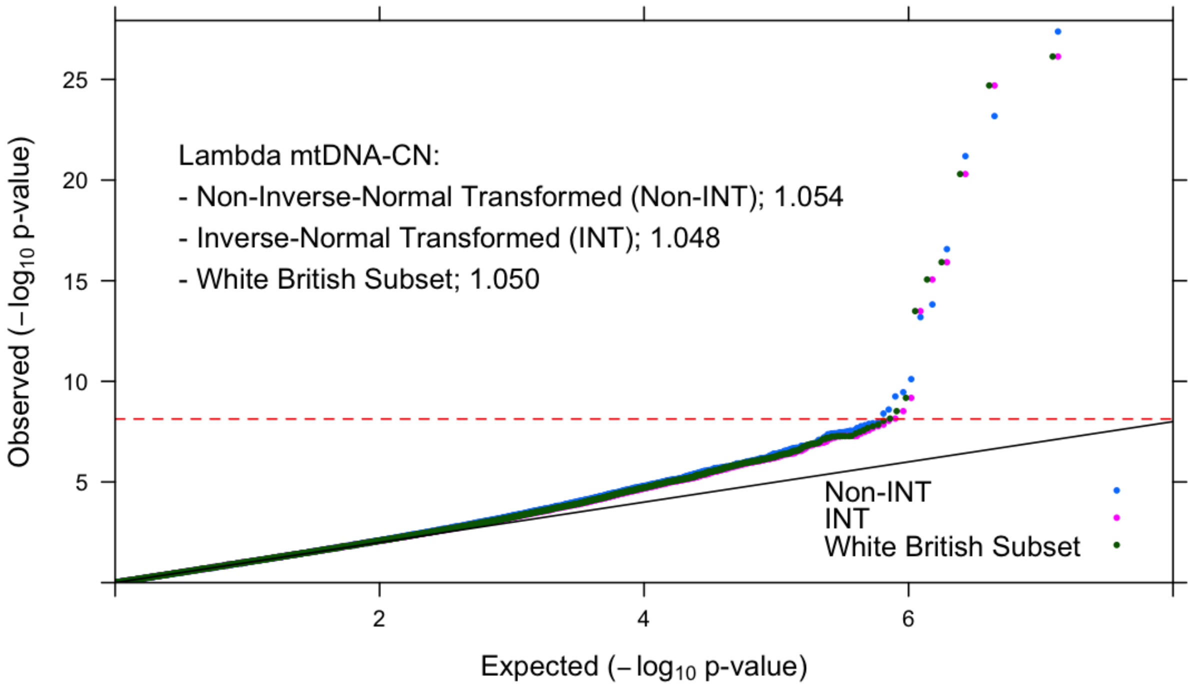

**Figure S7. mtDNA-CN by haplotype on 20q11.**

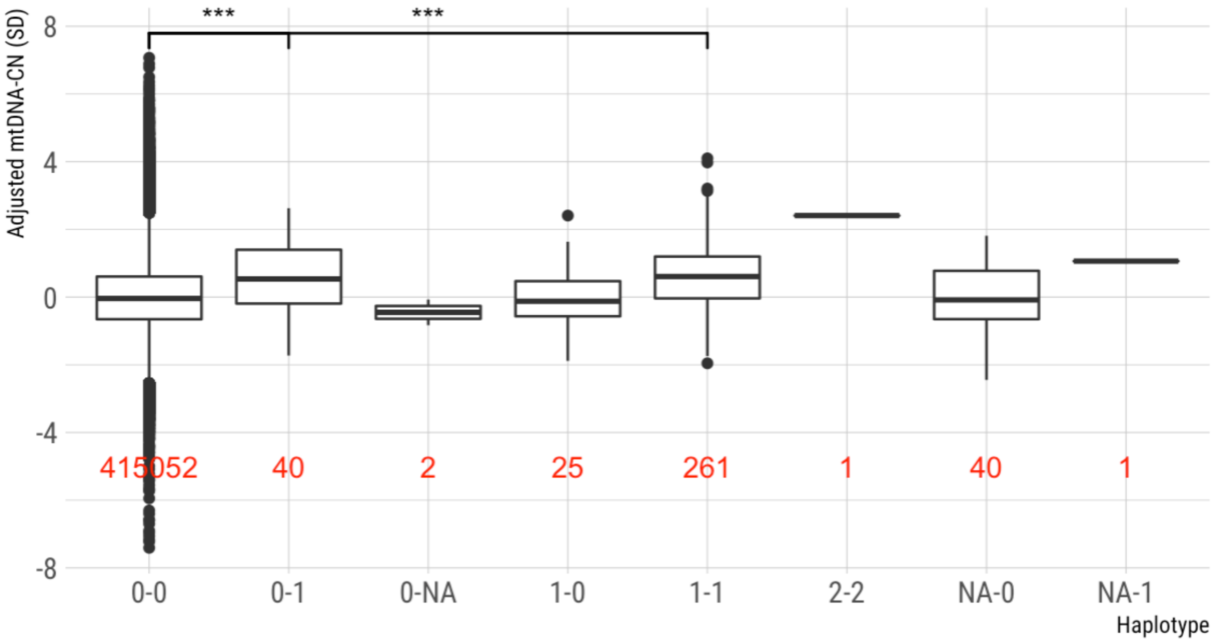


mtDNA-CN adjusted for age, sex, genetic ancestry PCs 1-40, and whole exome sequencing batch is plotted as a function of individuals harboring a specific haplotype on 20q11 in the UKB. **0-0** = 415,052 individuals with neither *SPAG4* nor *SAMHD1* variants (mean adjusted mtDNA-CN = 0 SD), **0-1** = 40 individuals with *SAMHD1* heterozygous variant only (mean adjusted mtDNA-CN = +0.6 SD), **1-0** = 25 individuals with *SPAG4* heterozygous variant only (mean adjusted mtDNA-CN = +0.01 SD), **1-1** = 261 individuals sharing both heterozygous variants in *SPAG4* and *SAMHD1* (mean adjusted mtDNA-CN = +0.63 SD), **2-2** = a single individual sharing homozygous variants at both sites (adjusted mtDNA-CN = +2.41 SD), **NA-0** = 40 individuals with missing variant in *SPAG4* (mean adjusted mtDNA-CN = -0.02 SD), **0-NA** = three individuals with missing variant in *SAMHD1* (mean adjusted mtDNA-CN = -0.45 SD), and **NA-1** = a single individual with missing variant in *SPAG4* and a heterozygous *SAMHD1* variant (adjusted mtDNA-CN = +1.06 SD). Stars represent p-value of t-test comparing adjusted mtDNA-CN between 0-0 and 0-1 (p-value = 0. 002) and between 0-0 and 1-1 (p-value = $1.94 \times 10^{-20}$). Not shown: t-test comparing adjusted mtDNA-CN between 0-0 and combined haplotypes of 1-1 and 2-2 (i.e. 262 carriers of both sites, p-value = $9.94 \times 10^{-21}$).

**Figure S8. Recombination rate on chromosome 20**

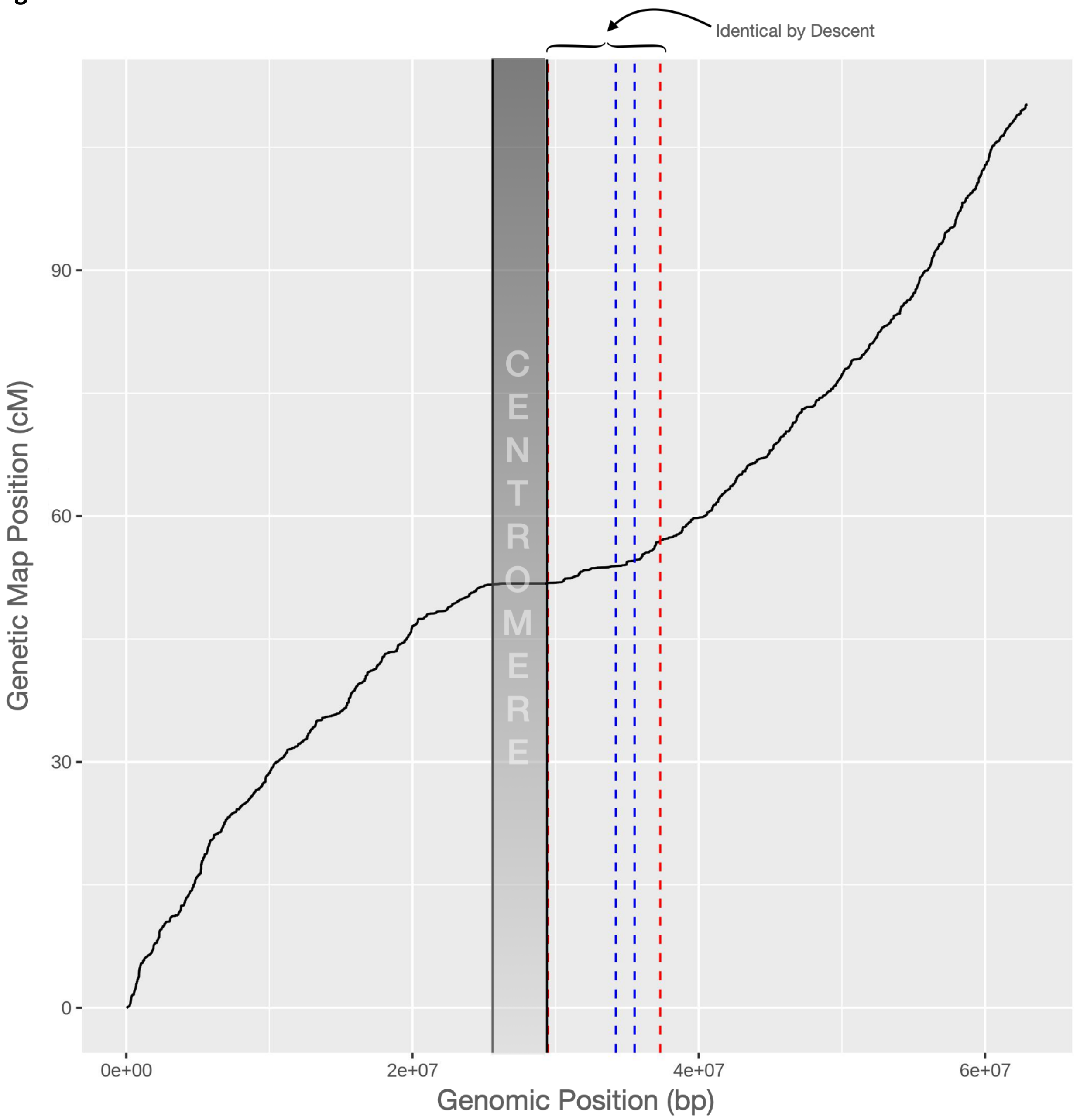


The physical positions on GRCh38 chromosome 20 are plotted against the genetic positions. Slope of line indicates recombination rate. Blue dashed lines indicate genomic coordinates of rare, shared haplotype between variants in *SPAG4* (hg38:20:35619007) and *SAMHD1* (hg38:20: 36893060). cM = centimorgan, bp = base pair.

### REFERENCES


1. Longchamps, R. J. *et al.* Genome-wide analysis of mitochondrial DNA copy number reveals loci implicated in nucleotide metabolism, platelet activation, and megakaryocyte proliferation. *Hum Genet* **141**, 127–146 (2022).
